# Pre-season screening currently has no value for injury prediction: The development and internal validation of a multivariable prognostic model to predict indirect muscle injury risk in elite football (soccer) players

**DOI:** 10.1101/19012054

**Authors:** Tom Hughes, Richard D. Riley, Michael J. Callaghan, Jamie C. Sergeant

**Affiliations:** Manchester United Football Club, AON Training Complex, Birch Road, Off Isherwood Road, Carrington, Manchester. UK; Centre for Epidemiology Versus Arthritis, Centre for Musculoskeletal Research, Manchester Academic Health Science Centre, University of Manchester, Manchester, UK; Centre for Prognosis Research, School of Primary, Community and Social Care, Keele University, Staffordshire, UK; Centre for Biostatistics, University of Manchester, Manchester Academic Health Science Centre, Manchester, UK; Department of Health Professions, Manchester Metropolitan University, Brooks Building, Bonsall Street, Manchester. UK

**Keywords:** Athlete, Injury prevention, Sport, Sprains and strains, Risk

## Abstract

**Background:** In elite football (soccer), periodic health examination (PHE) could provide prognostic factors to predict injury risk.

**Objective:** To develop and internally validate a prognostic model to predict individual indirect (non-contact) muscle injury (IMI) risk during a season in elite footballers, only using PHE-derived candidate prognostic factors.

**Methods:** Routinely collected preseason PHE and injury data were used from 119 players over 5 seasons (1^st^ July 2013 to 19^th^ May 2018). Ten candidate prognostic factors (12 parameters) were included in model development. Multiple imputation was used to handle missing values. The outcome was any time-loss, index indirect muscle injury (I-IMI) affecting the lower extremity. A full logistic regression model was fitted, and a parsimonious model developed using backward-selection to remove non-significant factors. Predictive performance was assessed through calibration, discrimination and decision-curve analysis, averaged across all imputed datasets. The model was internally validated using bootstrapping and adjusted for overfitting.

**Results:** During 317 participant-seasons, 138 I-IMIs were recorded. The parsimonious model included only age and frequency of previous IMIs; apparent calibration was perfect but discrimination was modest (C-index = 0.641, 95% confidence interval (CI): 0.580 to 0.703), with clinical utility evident between risk thresholds of 37-71%. After validation and overfitting adjustment, performance deteriorated (C-index = 0.580; calibration-in-the-large =-0.031, calibration slope =0.663).

**Conclusion:** The selected PHE data were insufficient prognostic factors from which to develop a useful model for predicting IMI risk in elite footballers. Further research should prioritise identifying novel prognostic factors to improve future risk prediction models in this field.

**Trial registration number:** NCT03782389

**KEY POINTS:**

- Factors measured through preseason screening generally have weak prognostic strength for future indirect muscle injuries and further research is needed to identify novel, robust prognostic factors.
- Because of sample size restrictions, and until the evidence base improves, it is likely that any further attempts at creating a prognostic model at individual club level would also suffer from poor performance.
- The value of using preseason screening data to make injury predictions or to select bespoke injury prevention strategies remains to be demonstrated, so screening should only be considered as useful for detection of salient pathology or for rehabilitation/ performance monitoring purposes at this time.

## 1. BACKGROUND

In elite football (soccer), indirect (non-contact) muscle injuries (IMIs) predominantly affect the lower extremities and account for 30.3% to 47.9% of all injuries that result in time lost to training or competition [1-5]. Reduced player availability negatively impacts upon medical [6] and financial resources [7, 8], and has implications for team performance [9]. Therefore, injury prevention strategies are important to professional teams [9].

Periodic health examination (PHE), or screening, is a key component of injury prevention practice in elite sport [10]. PHE is used by 94% of elite football teams and consists of medical, musculoskeletal, functional and performance tests, performed during preseason and in-season periods [11]. PHE has a rehabilitation and performance monitoring function [12] and is also used to detect musculoskeletal or medical conditions that may be dangerous or performance limiting [13]. Another perceived role of PHE is to recognise and manage factors that may increase, or predict, an athlete’s future injury risk [10], although this function is currently unsubstantiated [13].

PHE-derived variables associated with particular injury outcomes (such as IMIs) are called prognostic factors [14], which can be used to identify risk differences between players within a team [12]. Single prognostic factors are unlikely to satisfactorily predict an individual’s injury risk if used independently [15]. However, several factors could be combined in a multivariable prognostic prediction model to offer more accurate personalised risk estimates for the occurrence of a future event or injury [15, 16]. Such models could be used to identify high-risk individuals who may require an intervention that is designed to reduce risk [17], thus assisting clinical decision making [18]. Despite the potential benefits of prognostic models for injury, we are unaware of any that have been developed using PHE data in elite football [19].

Therefore, the aim of this study was to develop and internally validate a prognostic model to predict individualised IMI risk during a season in elite footballers, using PHE-derived candidate prognostic factors.

## 2. METHODS

The methods have been described in a published protocol [20] so will only be briefly outlined. This study has been registered on ClinicalTrials.gov (Identifier: NCT03782389) and is reported according to the Transparent Reporting of a Multivariable Prediction Model for Individual Prognosis or Diagnosis (TRIPOD) statement [21, 22].

### 2.1 Data Sources

This study was a retrospective cohort design. Eligible participants were identified from a population of male elite footballers, aged 16-40 years old at an English Premier League club. A dataset was created using routinely collected injury and preseason PHE data over 5 seasons (1^st^ July 2013 to 19^th^ May 2018). For each season (starting on 1^st^ July), participants completed a mandatory PHE during week 1 and were followed up to the final first team game of the season. If eligible participants were injured at the time of PHE, a risk assessment was completed by medical staff. Only tests that were appropriate and safe for the participant’s condition were completed; examiners were not blinded to injury status.

### 2.2 Participants and eligibility criteria

During any season, participants were eligible if they: 1) were not a goalkeeper; and 2) participated in PHE for the relevant season. Participants were excluded if they were not contracted to the club for the forthcoming season at the time of PHE.

### 2.3 Ethics and Data Use

Informed consent was not required as data were captured from the mandatory PHE completed through the participants’ employment. The data usage was approved by the club and University of Manchester Research Ethics Service.

### 2.4 Outcome

The outcome was any time-loss, index lower extremity IMI (I-IMI). That is, any I-IMI sustained by a participant during matches or training, which affected lower abdominal, hip, thigh, calf or foot muscle groups and prohibited future football participation [23]. I-IMIs were graded by a club doctor or physiotherapist (not blinded to PHE data) according to the validated Munich Consensus Statement for the Classification of Muscle Injuries in Sport [24, 25], during routine assessments undertaken within 24h of injury.

### 2.5 Sample size

We allowed a maximum of one candidate prognostic factor parameter per 10 I-IMIs, which (at the time of protocol development) was the main recommendation to minimise overfitting (Online Resource 1) [20]. The whole dataset was used for model development and internal validation, which agrees with methodological recommendations [26].

### 2.6 Candidate Prognostic Factors

The dataset contained 60 candidate factors [20]. Because of the sample size considerations, before any analysis, the set of candidates was reduced based upon data quality and reliability assessment, previous evidence of prognostic value [19] and clinical reasoning [20]. This process left a final set of 10 candidate factors, represented by 12 model parameters (Table 1).

**Table 1.** Set of candidate prognostic factors (with corresponding number of parameters) for model development.

| Selection method | Candidate Prognostic factor | Measurement unit | Number of model parameters corresponding to PF | Measurement method | Data type | Reliability (if applicable) |
| --- | --- | --- | --- | --- | --- | --- |
| Systematic review/clinical reasoning | Age | Years & days | 1 | Date of birth | Continuous | - |
|  | Frequency of previous muscle injuries within 3 years prior to PHE | Count | 1 | Medical records | Discrete (treated as continuous) | - |
|  | Most recent previous muscle injury within 3 years prior to baseline PHE | Never (ref); < 6 months; 6-12 months; > 12 months | 3 | Medical records | Categorical | - |
| Data quality/clinical reasoning | CMJ peak power | Watts | 1 | CMJ using force plates | Continuous | Test-retest ICC = 0.92-0.98 [27] |
|  | PROM hip joint internal rotation difference* | Degrees | 1 | Digital inclinometer | Continuous | Intra-rater ICC = 0.90 [28] |
|  | PROM hip joint external rotation difference* | Degrees | 1 | Digital inclinometer | Continuous | Intra-rater ICC = 0.90 [28] |
|  | Hip flexor muscle length difference* | Degrees | 1 | Thomas test using digital inclinometer | Continuous | Inter-rater ICC = 0.89 [29] |
|  | Hamstring muscle length/neural mobility difference* | Degrees | 1 | SLR using digital inclinometer | Continuous | Intra-rater ICC = 0.95-0.98 [30]<br>Interrater ICC = 0.80-0.97 [31] |
|  | Calf muscle length difference* | Degrees | 1 | WBL using digital inclinometer | Continuous | Inter-rater ICC = 0.80- 0.95 [32, 33] Intra-rater = ICC 0.88 [33] |
|  | BMI | Kg/m<sup>2</sup> | 1 | Composite height (cm) and weight (kg) | Continuous | - |

Key: PF= prognostic factor; PHE=periodic health examination; ref= reference category (does not count as a model parameter); WBL=weight bearing lunge;
CMJ=countermovement jump; PROM=passive range of movement; ICC=intraclass correlation coefficient; SLR= straight leg raise; BMI= body mass index; Kg=kilos; m = mass; - = not applicable; Note: \* denotes between limb differences.

## 3. Statistical analysis

### 3.1 Data handling – outcome measures

Each participant-season was treated as independent. Participants who sustained an an I-IMI were no longer considered at risk for that season and were included for further analysis at the start of the next season if still eligible. Any upper limb IMI, trunk IMI or non-IMI injuries were ignored and participants were still considered at risk.

Eligible participants who were loaned to another club throughout that season, but had not sustained an I-IMI prior to the loan were still considered at risk. I-IMIs that occurred whilst on loan were included for analysis, as above. Permanently transferred participants (who had not sustained an I-IMI prior to leaving), were recorded as not having an I-IMI during the relevant season and exited the cohort at the season end.

### 3.2 Data handling –missing data

Missing values were assumed to be missing at random [20]. The continuous parameters generally demonstrated non-normal distributions, so were transformed using normal scores [34] to approximate normality before imputation, and back-transformed following imputation [35]. Multivariate normal multiple imputation was performed, using a model that included all candidates and I-IMI outcomes. Fifty imputed datasets were created in Stata 15.1 (StataCorp LLC, Texas, USA) and analysed using the ‘*mim*’ module.

### 3.3 Prognostic Model Development

Continuous parameters were retained on their original scales and their effects assumed linear [22]. A full multivariable logistic regression model was constructed, which contained all 12 parameters. Parameter estimates and results were combined across imputed datasets using Rubin’s Rules [36]. To develop a parsimonious model that would be easier to utilise in practice, backward variable selection was performed simultaneously across all imputed datasets to successively remove non-significant factors with p-values > 0.157 (approximate equivalence with Akaike’s Information Criterion) [37, 38]. Multiple parameters representing the same candidate factor were tested together so that the whole factor was either retained or removed. Candidate interactions were not examined and no terms were forced into the model. All analyses were conducted in Stata 15.1.

### 3.4 Assessment of model performance

The parsimonious model was used to predict I-IMI risk over a season, for every participant-season in all imputed datasets. For each performance measure, the model’s apparent performance was assessed in each imputed dataset and then averaged across all imputed datasets using Rubin’s Rules [36]. Discrimination determines a model’s ability to differentiate between participants who have experienced an outcome compared to those who have not [39], quantified using the concordance index (c-index). This is equivalent to the area under the receiver operating characteristic (ROC) curve for logistic regression, where 1 demonstrates perfect discrimination, while 0.5 indicates that discrimination is no better than chance [40].

Calibration determines the agreement between the model’s predicted outcome risks and those observed [41], evaluated using a calibration plot in each imputed dataset. All predicted risks were divided into ten groups defined by tenths of predicted risk. The mean predicted risks for the groups were plotted against the observed group outcome proportions with corresponding 95% confidence intervals (CIs). A loess smoothing algorithm showed calibration across the range of predicted values [42]. For grouped and smoothed data points, perfect predictions lie on the 45° line (i.e. a slope of 1).

The systematic (mean) error in model predictions was quantified using calibration-in-the-large (CITL), which has an ideal value of 0 [39, 41], and the expected/observed (E/O) statistic, which is the ratio of the mean predicted risk against the mean observed risk (ideal value of 1) [39, 41]. The degree of over or underfitting was determined using the calibration slope, where a value of 1 equals perfect calibration on average across the entire range of predicted risks [22]. Nagelkerke’s pseudo-R^2^ was also calculated, which quantifies the overall model fit, with a range of 0 (no variation explained) to 1 (all variation explained) [43].

### 3.5 Assessment of clinical utility

Decision-curve analysis was used to assess the model’s clinical usefulness in terms of net benefit (NB) if used to allocate possible preventative interventions. This assumed that the model’s predicted risks were classed as positive (i.e. may require a preventative intervention) if greater than a chosen risk threshold, and negative otherwise. NB is then the difference between the proportion of true positives and false positives (weighted by the odds of the chosen risk threshold), divided by the sample size [44]. Positive NB values suggest the model is beneficial compared to treating none (which has no benefit to the team but with no negative cost and efficiency implications). The maximum possible NB value is the proportion with the outcome in the dataset.

The model’s NB was also compared to the NB of delivering an intervention to all individuals (a treat-all strategy, offering maximum benefit to the team, but with maximum negative cost and efficiency implications) [17]. A model has potential clinical value if it demonstrates higher NB than the default strategies over the range of risk thresholds which could be considered as high risk in practice [45].

### 3.6 Internal validation and adjustment for overfitting

To examine overfitting, the model was internally validated using 200 bootstrap samples, drawn from the original dataset. In each sample, the complete model-building procedure (including multiple imputation, backward variable selection and performance assessment) was conducted as described earlier. The difference in apparent performance (of a bootstrap model in its bootstrap sample) and test performance (of the bootstrap model in the original dataset) was averaged across all samples. This generated optimism estimates for the calibration slope, CITL and C-index statistics. These were subtracted from the original apparent calibration slope, CITL and C-index statistics to obtain final optimism-adjusted performance estimates.

To produce a final model adjusted for overfitting, the regression coefficients produced in the parsimonious model were multiplied by the optimism-adjusted calibration slope (uniform shrinkage factor) to adjust (shrink) for overfitting [46]. Finally, the CITL (model intercept) was then re-estimated to give the final model, suitable for evaluation in other populations or datasets.

### 3.7 Complete case and sensitivity analyses

To determine the effect of multiple imputation and player transfer assumptions on model stability, the model development process was repeated: 1) as a complete case analysis; and 2) as sensitivity analyses excluding participant-seasons for participants who were loaned or transferred (performed as both multiple imputation and complete case analyses).

## 4. RESULTS

### 4.1 Participants

During the five seasons, 119 participants were included, contributing 317 participant-seasons and 138 IMIs in the primary analyses (Fig. 1). For the sensitivity analyses (excluding loans and transfers), 265 independent participant-seasons with 120 IMIs were included; 47 participants were transferred during a season, which excluded 52 participant-seasons (Fig. 1).

**Fig 1.**
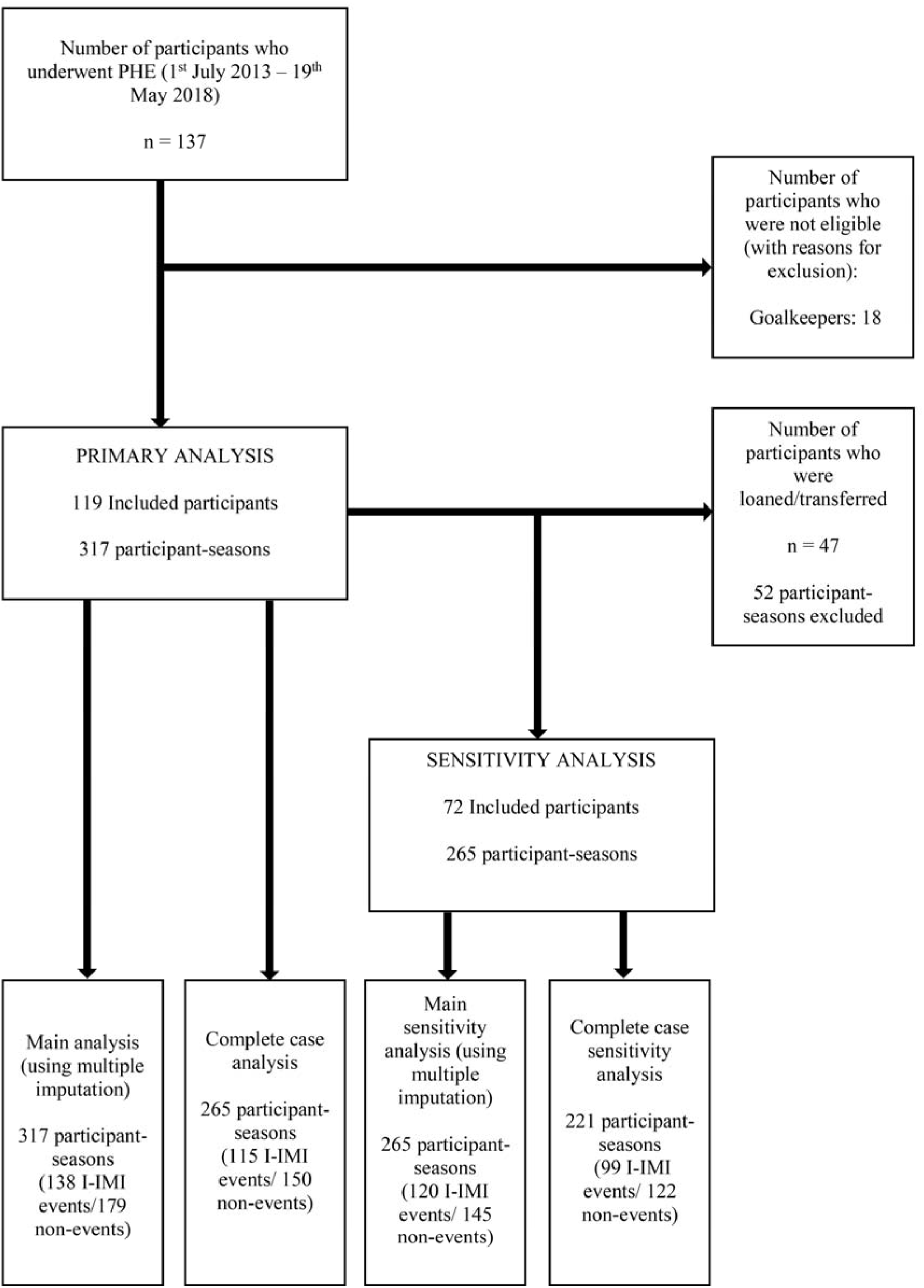
Participant flow chart *Key: n=participants; I-IMI=index indirect muscle injury*.

Table 2 shows anthropometric and all prognostic factor characteristics for participants included in the primary analyses. These were similar to those included in the sensitivity analyses (Online Resource 2).

**Table 2.** Characteristics of included participants

| *Characteristic/candidate prognostic factors* | *Measurement method* | *Data type* | *Freq. (%) if categorical* | *Min* | *Lower quartile* | *Median* | *Upper quartile* | *Max* | *Missing values n (%)* |
| --- | --- | --- | --- | --- | --- | --- | --- | --- | --- |
| **Anthropometrics** |  |  |  |  |  |  |  |  |  |
| Age at PHE (years) | Birthdate | Cont. | - | 16.01 | 17.80 | 19.69 | 23.56 | 39.59 | 0 (0) |
| Height (cm) | Standing height | Cont. | - | 164.3 | 176.0 | 180.0 | 185.5 | 195.0 | 18 (5.68) |
| Weight (kg) | Digital scales | Cont. | - | 56.8 | 69.2 | 73.6 | 80.0 | 94.0 | 18 (5.68) |
| BMI (Kg/m<sup>2</sup>) | Calculated using the formula: Kg/m<sup>2</sup> | Cont. | - | 18.1 | 21.8 | 22.7 | 23.7 | 29.1 | 23 (7.26) |
| **Past medical history** |  |  |  |  |  |  |  |  |  |
| Freq. of previous IMIs in 3 years prior to PHE | Medical records | Dis./cont. | - | 0 | 0 | 1 | 2 | 7 | 0 (0) |
| Most recent previous IMI in 3 years prior to PHE |  |  |  |  |  |  |  |  |  |
| Never | Medical records | Cat. | 143 (45.11) | - | - | - | - | - | 0 (0) |
| <6 months | Medical records | Cat. | 48 (15.14) | - | - | - | - | - | 0 (0) |
| 6-12 months | Medical records | Cat. | 52 (16.40) | - | - | - | - | - | 0 (0) |
| >12 months | Medical records | Cat. | 74 (23.34) | - | - | - | - | - | 0 (0) |
| **Musculoskeletal Examination** |  |  |  |  |  |  |  |  |  |
| PROM hip internal rotation difference (deg.) | Hip PROM with inclinometer | Cont. | - | -25.0 | -3.0 | 0.0 | 5.0 | 20.0 | 20 (6.31) |
| PROM hip external rotation difference (deg.) | Hip PROM with inclinometer | Cont. | - | -20.0 | -5.0 | 0.0 | 5.0 | 25.0 | 20 (6.31) |
| Hip flexor length difference (deg.) | Thomas test with inclinometer | Cont. | - | -20.0 | -2.0 | 0.0 | 3.0 | 14.0 | 23 (7.26) |
| Hamstring length /neural mobility difference (deg.) | SLR with inclinometer | Cont. | - | -20.0 | 0.0 | 0.0 | 0.0 | 15.0 | 23 (7.26) |
| Calf muscle length difference (deg.) | WBL with inclinometer | Cont. | - | -20.0 | -2.0 | 0.0 | 3.0 | 15.0 | 20 (6.31) |
| **Lower Extremity Power** |  |  |  |  |  |  |  |  |  |
| CMJ power (Watts) | CMJ using force platform | Cont. | - | 2625.0 | 3707.0 | 4150.0 | 4662.0 | 6577.0 | 42 (13.25) |

*Key: PHE= periodic health examination; I-IMI=index indirect muscle injury; IMI= indirect muscle injury; min = minimum; max = maximum; n = observations; Freq= frequency;*
*WBL=weight bearing lunge; CMJ=countermovement jump; PROM=passive range of movement; deg. = degrees; SLR= straight leg raise; BMI= body mass index; kg/m<sup>2</sup> = kilograms/body height (metres) squared; cm = centimetres; Kg=kilograms; Cont.=continuous; dis./cont. = discrete treated as continuous; cat. = categorical.*

### 4.2 Missing data and multiple imputation

All I-IMI, age and previous muscle injury data were complete (Table 2). For all other candidates, missing data ranged from 6.31% (for hip internal and external rotation difference) to 13.25% for countermovement jump (CMJ) power (Table 2). The distribution of imputed values approximated observed values (Online Resource 3), confirming their plausibility.

### 4.3 Model development

Table 3 shows the parameter estimates for the full model and parsimonious model after variable selection (averaged across imputations).

**Table 3.**
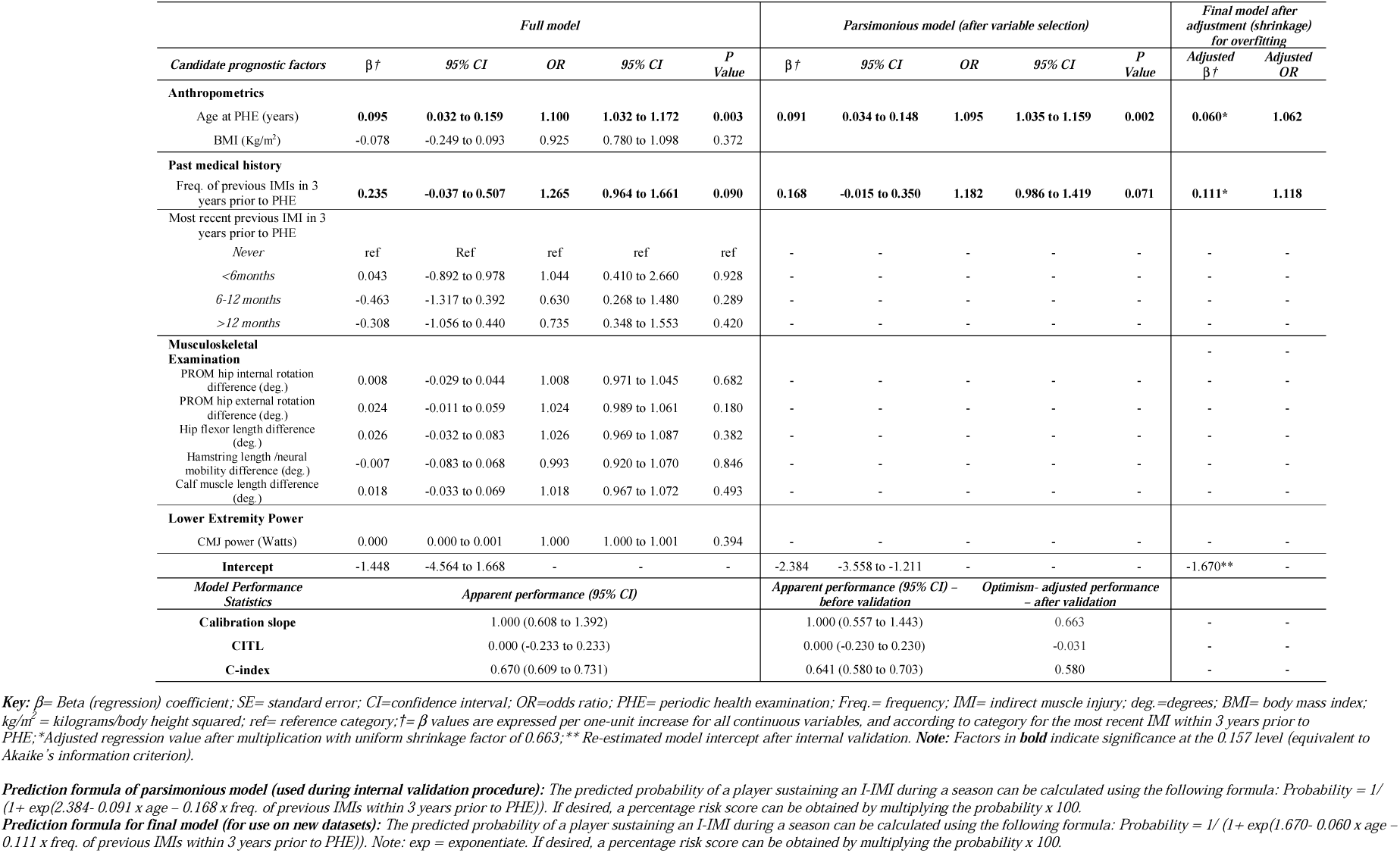
Results of the full multivariable logistic regression model, the parsimonious model after variable selection and the formula to derive individual predictions – Primary analysis using imputed data

For both models, only age and frequency of previous IMIs had a statistically significant (but modest) association with increased I-IMI risk (p <0.157). No clear evidence for an association was observed for any other candidate factor.

### 4.4 Model performance assessment and clinical utility

Table 3 shows the apparent performance measures for the full and parsimonious models. Fig. 2 shows a calibration plot for the parsimonious model. These were identical across all imputed datasets because the retained prognostic factors contained no missing values. The parsimonious model had perfect apparent overall CITL and calibration slope by definition, but calibration was more variable around the 45° degree line between the expected risk ranges of 28% to 54%. Discrimination was similarly modest for the full (C-index= 0.670, 95% CI=0.609 to 0.731) and parsimonious models (C-index = 0.641, 95%CI = 0.580-0.703). The overall model fit after variable selection was low (Nagelkerke R^2^ = 0.089).

**Fig. 2.**
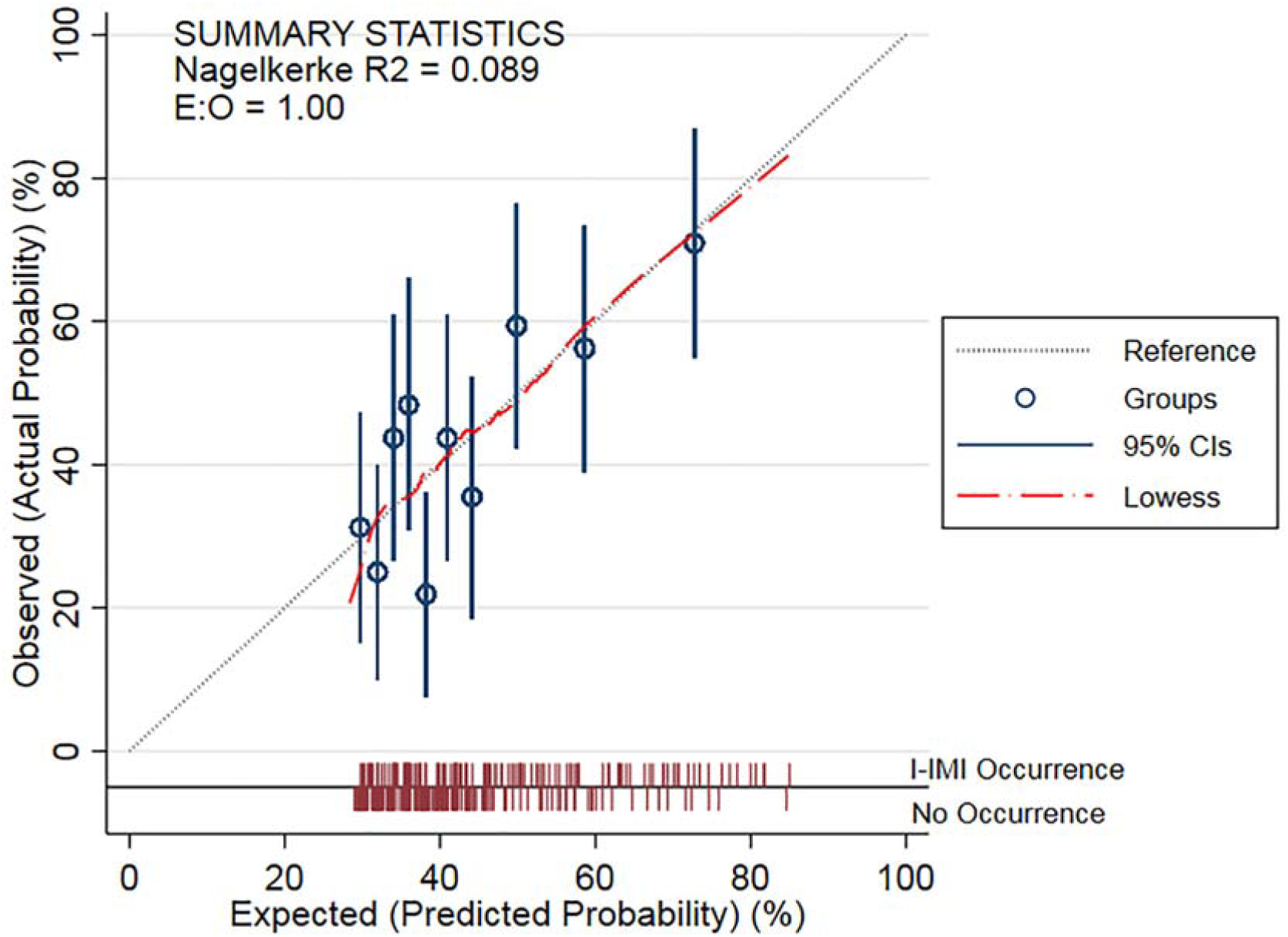
Calibration plot for the parsimonious prognostic model (before adjustment for overfitting), showing apparent calibration in the dataset used to develop the model. Primary analysis using one of the imputed datasets Key: E:O= expected:observed ratio; CI= confidence interval.

Fig. 3 displays the decision-curve analysis. The NB of the parsimonious model was comparable to the treat-all strategy at risk thresholds up to 31%, marginally greater between 32% and 36% and exceeded the NB of either default strategies between 37% and 71%.

**Fig. 3.**
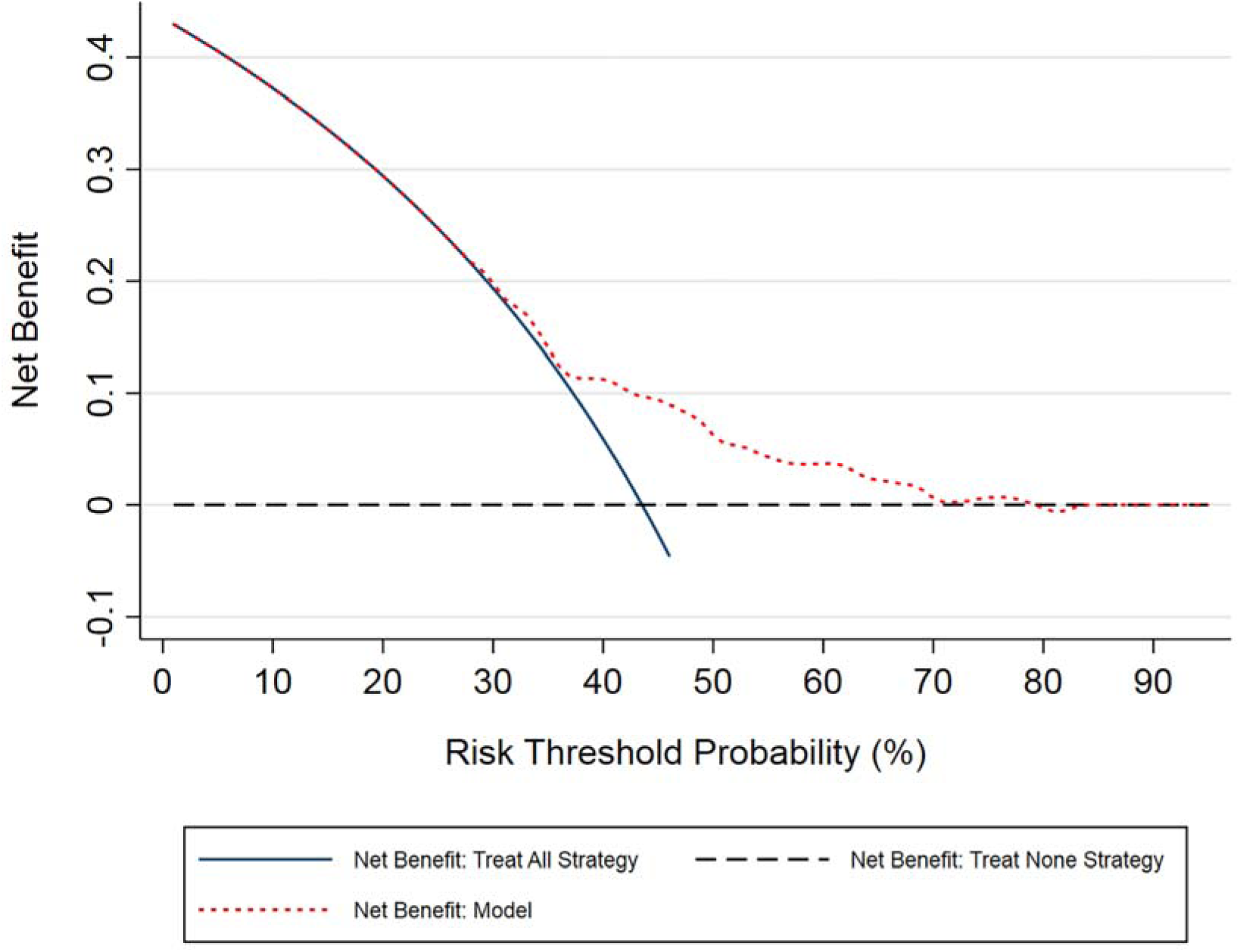
Decision curve plot for the parsimonious model (before adjustment for overfitting) compared to the treat all or treat none default strategies.

### 4.5 Complete Case and Sensitivity Analyses

The full and parsimonious models were robust to complete case analyses and excluding loans and transfers, with comparable performance estimates (c-index range= 0.632-0.678; Nagelkerke R^2^ range= 0.102 to 0.130) (Online Resources 4-7). The same prognostic factors were selected in all parsimonious models.

### 4.6 Internal validation and adjustment for overfitting

Table 3 shows the optimism-adjusted performance statistics for the parsimonious model, with full internal validation results shown in Online Resource 8. After adjustment for optimism, the model’s discrimination performance deteriorated (c-index = 0.580). Furthermore, bootstrapping suggested the model would be severely overfitted in new data (calibration slope = 0.663), so a shrinkage factor of 0.663 was applied to the parsimonious parameter estimates and the model intercept re-estimated to produce our final model (Table 3).

## 5. DISCUSSION

We have developed and internally validated a multivariable prognostic model to predict individualised I-IMI risk during a season in elite footballers, only using retrospectively collected preseason PHE and injury data. This is the only study that we know of that has developed a prognostic model for this purpose, so the results cannot be compared to previous work.

The performance of the full and parsimonious models was similar, which means that utilising all candidate factors offered very little advantage over using two for making predictions. Indeed, variable selection eliminated 8 candidate prognostic factors that had no clear evidence for an association with I-IMIs. Our findings confirm previous suggestions that PHE tests designed to measure modifiable physical and performance characteristics typically offer poor predictive value [10]. This may be because unless particularly strong associations are observed between a PHE test and injury outcome, the overlap in scores between individuals who sustain a future injury and those who do not results in poor discrimination [10]. Additionally, after measurement at a single timepoint (i.e. pre-season), it is likely that the prognostic value of these modifiable factors may vary over time [47] due to training exposure, environmental adaptations and the occurrence of injuries [48].

The variable selection process resulted in a model which included only age and the frequency of previous IMIs within the last three years, which are simple to measure and routinely available in practice. Our findings were similar to the modest association previously observed between age and hamstring IMIs in elite players [19]. However, while a positive previous hamstring IMI history has a confirmed association with future hamstring IMIs [19], we found that for lower extremity I-IMIs, cumulative IMI frequency was preferred to the time proximity of any previous IMI as a multivariable prognostic factor. Nevertheless, the weak prognostic strength of these factors explains the parsimonious model’s poor discrimination and low potential for clinical utility.

Our study is the first to examine the clinical usefulness (net benefit) of a model to identify players at high risk of IMIs and who may benefit from preventative interventions such as training load management, strength and conditioning or physiotherapy programmes. Our parsimonious model demonstrated no clinical value at risk thresholds of less than 36%, because its NB was comparable to that of providing all players with an intervention. Indeed, the only clinically useful thresholds that would indicate a high-risk player would be 37-71%, where the model’s NB was greater than giving all players an intervention. However, because of the high baseline IMI risk in our population (approximately 44% of participant-seasons affected), the burden of IMIs [1-5] and the minimal costs [10] versus the potential benefits of such preventative interventions in an elite club setting, these thresholds are likely to be too high to be acceptable in practice. Accordingly, it would be inappropriate to allocate or withhold interventions based upon our model’s predictions.

Because of severe overfitting our parsimonious model was optimistic, which means that if used in new players, prediction performance would likely to be worse [38]. Although our model was adjusted (shrunk) to account for overfitting and hence improve its calibration performance in new datasets, given the limitations in performance and clinical value, we cannot recommend that it is validated externally or used in clinical practice.

This study has some limitations. We measured candidate factors at one timepoint each season and assumed that participant-seasons were independent. While statistically complex, future studies may improve predictive performance and external validity by harnessing longitudinal measurements and incorporating between-season correlations.

We also merged all lower extremity I-IMIs rather than using specific muscle group outcomes. Although less clinically meaningful, this was necessary to maximise statistical power. Nevertheless, our limited sample size prohibited examination of complex non-linear associations and permitted a small number of candidates to be considered. A lack of known prognostic factors [19] meant that selection was mainly guided by data quality control processes and clinical reasoning, so it is possible that important factors were not included. Risk prediction improves when multiple factors with strong prognostic value are used [15]. Therefore, future research should aim to identify novel prognostic factors, so that these can be used to develop models with greater potential clinical benefit. This may also allow updating of our model to improve its performance and clinical utility [49].

Until the evidence base improves, and because of sample size limitations, it is likely that any further attempts to create a prognostic model at individual club level would suffer similar issues. Importantly, this means that for any team, the value of using preseason PHE data to make individualised predictions or to select bespoke injury prevention strategies remains to be demonstrated. However, the pooling of individual participant data from several participating clubs may increase sample sizes sufficiently to allow further model development studies [50], where a greater number of candidate factors could be utilised.

## 6. CONCLUSION

Using PHE and injury data available pre-season, we have developed and internally validated a prognostic model to predict I-IMI risk in players at an elite club, using current methodological best practice. The paucity of known prognostic factors and data requirements for model building severely limited the model’s performance and clinical utility, so it cannot be recommended for external validation or use in practice. Further research should prioritise identifying novel prognostic factors to improve future risk prediction models in this field.

## Data Availability

An anonymised summary of the dataset that was analysed during this study may be available from the corresponding author on reasonable request.

## CONTRIBUTORS

TH was responsible for the conceptualisation of the project, study design, database construction, data extraction and cleaning, protocol development and protocol writing. TH conducted the data analysis, interpretation and wrote the main manuscript. RR provided statistical guidance and assisted with development of the study design, analysis, and edited manuscript drafts. MC assisted with the study conceptualisation and design, protocol development, clinical interpretation and editing the manuscript drafts. JS provided guidance with the study design, development of the analysis and protocol, supervision and interpretation of the analysis, as well as editing the study manuscripts. All authors read and approved this final manuscript.

## COMPLIANCE WITH ETHICAL STANDARDS

### Funding

The lead researcher (TH) is receiving sponsorship from Manchester United Football Club to complete a postgraduate PhD study programme. This work was also supported by Versus Arthritis: grant number 21755.

### Informed consent

Informed consent was not required as data were captured from the mandatory PHE completed through the participants’ employment.

### Ethical approval

The data usage was approved by the football club and the Research Ethics Service at the University of Manchester.

## PATIENT CONSENT FOR PUBLICATION

Not required.

## ACKNOWLEDGEMENTS

The authors would like to thank all staff within the Medical and Sports Science Department at Manchester United for their continuing help and support with this manuscript and thank all players for their participation (without whom this study would not be possible). The authors also thank the Centre for Epidemiology Versus Arthritis for their support: Versus Arthritis grant number 21755.

## REFERENCES

1. Ekstrand J, Hagglund M, Walden M. Epidemiology of muscle injuries in professional football (soccer). Am J Sports Med. 2011 Jun;39(6):1226–32.

2. Falese L, Della Valle P, Federico B. Epidemiology of football (soccer) injuries in the 2012/2013 and 2013/2014 seasons of the Italian Serie A. Res Sports Med. 2016;24(4):426–32.

3. Larruskain J, Lekue JA, Diaz N, Odriozola A, Gil SM. A comparison of injuries in elite male and female football players: A five-season prospective study. Scand J Med Sci Sports. 2018;28(1):237–45.

4. Leventer L, Eek F, Hofstetter S, Lames M. Injury Patterns among Elite Football Players: A Media-based Analysis over 6 Seasons with Emphasis on Playing Position. Int J Sports Med. 2016;37(11):898–908.

5. Hawkins RD, Fuller CW. A prospective epidemiological study of injuries in four English professional football clubs. Br J Sports Med. 1999;33(3):196–203.

6. Woods C, Hawkins R, Hulse M, Hodson A. The Football Association Medical Research Programme: an audit of injuries in professional football-analysis of preseason injuries. Br J Sports Med. 2002 Dec;36(6):436–41.

7. Ekstrand J. Preventing injuries in professional football: thinking bigger and working together. Br J Sports Med. 2016;50(12):709–10.

8. Ekstrand J. Keeping your top players on the pitch: the key to football medicine at a professional level. Br J Sports Med. 2013;47(12):723–4.

9. Hagglund M, Walden M, Magnusson H, Kristenson K, Bengtsson H, Ekstrand J. Injuries affect team performance negatively in professional football: an 11-year follow-up of the UEFA Champions League injury study. Br J Sports Med. 2013 Aug;47(12):738–42.

10. Bahr R. Why screening tests to predict injury do not work-and probably never will…: a critical review. Br J Sports Med. 2016 Jul;50(13):776–80.

11. McCall A, Carling C, Davison M, Nedelec M, Le Gall F, Berthoin S, et al. Injury risk factors, screening tests and preventative strategies: a systematic review of the evidence that underpins the perceptions and practices of 44 football (soccer) teams from various premier leagues. Br J Sports Med. 2015;49(9):583–9.

12. Hughes T, Sergeant JC, van der Windt DA, Riley R, Callaghan MJ. Periodic Health Examination and Injury Prediction in Professional Football (Soccer): Theoretically, the Prognosis is Good. Sports Med. 2018;48(11):2443–48.

13. Ljungqvist A, Jenoure PJ, Engebretsen AH, Alonso JM, Bahr R, Clough AF, et al. The International Olympic Committee (IOC) Consensus Statement on Periodic Health Evaluation of Elite Athletes. Clin J Sport Med. 2009;19(5):347–60.

14. Riley RD, van der Windt DA, Croft P, Moons KG. Prognosis Research in Healthcare: Concepts, Methods and Impact. Oxford: Oxford University Press; 2019.

15. Riley RD, Hayden JA, Steyerberg EW, Moons KG, Abrams K, Kyzas PA, et al. Prognosis Research Strategy (PROGRESS) 2: prognostic factor research. PLoS Med. 2013;10(2):e1001380.

16. Steyerberg EW, Moons KG, van der Windt DA, Hayden JA, Perel P, Schroter S, et al. Prognosis Research Strategy (PROGRESS) 3: prognostic model research. PLoS Med. 2013;10(2):e1001381.

17. Localio AR. Beyond the Usual Prediction Accuracy Metrics: Reporting Results for Clinical Decision Making. Ann Intern Med. 2012;157(4):294–6.

18. Bernard A. Clinical prediction models: a fashion or a necessity in medicine? J Thorac Dis. 2017;9(10):3456–7.

19. Hughes T, Sergeant JC, Parkes M, Callaghan MJ. Prognostic factors for specific lower extremity and spinal musculoskeletal injuries identified through medical screening and training load monitoring in professional football (soccer): a systematic review.BMJ Open Sport & Exercise Medicine. 2017;3(1):1–18.

20. Hughes T, Riley R, Sergeant J, Callaghan M. A study protocol for the development and internal validation of a multivariable prognostic model to determine lower extremity muscle injury risk in elite football (soccer) players, with further exploration of prognostic factors. Diagn Progn Res. 2019;3:19. https://doi.org/10.1186/s41512-019-0063-8

21. Collins GS, Reitsma JB, Altman DG, Moons KG. Transparent reporting of a multivariable prediction model for individual prognosis or diagnosis (TRIPOD): the TRIPOD statement. BMJ. 2015;350:g7594.

22. Moons KG, Altman DG, Reitsma JB, Ioannidis JP, Macaskill P, Steyerberg EW, et al. Transparent Reporting of a multivariable prediction model for Individual Prognosis or Diagnosis (TRIPOD): explanation and elaboration. Ann Intern Med. 2015;162(1):W1–73.

23. Fuller CW, Ekstrand J, Junge A, Andersen TE, Bahr R, Dvorak J, et al. Consensus statement on injury definitions and data collection procedures in studies of football (soccer) injuries. Br J Sports Med. 2006;40(3):193–201.

24. Mueller-Wohlfahrt HW, Haensel L, Mithoefer K, Ekstrand J, English B, McNally S, et al. Terminology and classification of muscle injuries in sport: the Munich consensus statement. Br J Sports Med. 2013;47(6):342–50.

25. Ekstrand J, Askling C, Magnusson H, Mithoefer K. Return to play after thigh muscle injury in elite football players: implementation and validation of the Munich muscle injury classification. Br J Sports Med. 2013;47(12):769–74.

26. Steyerberg EW, Uno H, Ioannidis JPA, van Calster B, Collaborators. Poor performance of clinical prediction models: the harm of commonly applied methods. J Clin Epidemiol. 2018;98:133–43.

27. Hori N, Newton RU, Kawamori N, McGuigan MR, Kraemer WJ, Nosaka K. Reliability of performance measurements derived from ground reaction force data during countermovement jump and the influence of sampling frequency. J Strength Cond Res. 2009;23(3):874–82.

28. Roach S, San Juan JG, Suprak DN, Lyda MA. Concurrent validity of digital inclinometer and universal goniometer assessing passive hip mobility in healthy subjects. Int J Sports Phys Ther. 2013;8(5):680–8.

29. Clapis PA, Davis SM, Davis RO. Reliability of inclinometer and goniometric measurements of hip extension flexibility using the modified Thomas test. Phys Theory Pract. 2008;24(2):135–41.

30. Boyd BS. Measurement properties of a hand-held inclinometer during straight leg raise neurodynamic testing. Physiotherapy. 2012;98(2):174–9.

31. Gabbe BJ, Bennell KL, Wajswelner H, Finch CF. Reliability of common lower extremity musculoskeletal screening tests. Phys Ther Sport. 2004;5(2):90–7.

32. Williams CM, Caserta AJ, Haines TP. The TiltMeter app is a novel and accurate measurement tool for the weight bearing lunge test. J Sci Med Sport. 2013;16(5):392–5.

33. Munteanu SE, Strawhorn AB, Landorf KB, Bird AR, Murley GS. A weightbearing technique for the measurement of ankle joint dorsiflexion with the knee extended is reliable. J Sci Med Sport. 2009;12(1):54–9.

34. Lunt M. nscore. 2007 [cited; Available from: http://personalpages.manchester.ac.uk/staff/mark.lunt]

35. Sterne JAC, White IR, Carlin J, Spratt M, Royston P, Kenward MG, et al. Multiple imputation for missing data in epidemiological and clinical research: potential and pitfalls. Br Med J. 2009;338:b2393.

36. Marshall A, Altman DG, Holder RL, Royston P. Combining estimates of interest in prognostic modelling studies after multiple imputation: current practice and guidelines. BMC Med Res Methodol. 2009; 28; 9:57.

37. Sauerbrei W. The use of resampling methods to simplify regression models in medical statistics. J Royal Stat Soc. 1999;48(3):313–29.

38. Steyerberg EW, Eijkenmans MJ, Harrell FE, Habbema JDF. Prognostic Modelling with Logistic Regression Analysis: In search of a Sensible Strategy in Small Data Sets. Med Decis Making. 2001;21(1):45–56.

39. Steyerberg EW, Vergouwe Y. Towards better clinical prediction models: seven steps for development and an ABCD for validation. Eur Heart J. 2014;35(29):1925–31.

40. Royston P, Moons KG, Altman DG, Vergouwe Y. Prognosis and Prognostic Research:Developing a Prognostic Model Br Med J. 2009;338.

41. Steyerberg EW, Vickers AJ, Cook NR, Gerds T, Gonen M, Obuchowski N, et al. Assessing the Performance of Prediction Models: a framework for some traditional and novel measures. Epidemiology. 2010;21(1):128–38.

42. Austin PC, Steyerberg EW. Graphical assessment of internal and external calibration of logistic regression models by using loess smoothers. Stat Med. 2014 Feb 10;33(3):517–35.

43. Bewick V, Cheek L, Ball J. Statistics review 14: Logistic regression. Crit Care. 2005 Feb;9(1):112–8.

44. Van Calster B, Wynants L, Verbeek JFM, Verbakel JY, Christodoulou E, Vickers AJ, et al. Reporting and Interpreting Decision Curve Analysis: A Guide for Investigators. Eur Urol. 2018 Dec;74(6):796–804.

45. Vickers AJ, Van Calster B, Steyerberg EW. Net benefit approaches to the evaluation of prediction models, molecular markers, and diagnostic tests. BMJ. 2016;352:i6.

46. Harrell FE. Regression modelling strategies: with applications to linear models, logistic and ordinal regression, and survival analysis. 2nd ed. New York: Springer; 2015.

47. Bansal A, Heagerty PJ. A comparison of landmark methods and time-dependent ROC methods to evaluate the time-varying performance of prognostic markers for survival outcomes. Diagn Progn Res. 2019;3:14.

48. Meeuwisse W, Tyreman H, Hagel B, Emery C. A Dynamic Model of Etiology in Sport Injury: The Recursive Nature of Risk and Causation. Clin J Sports Med. 2007;17(3):215–9.

49. Steyerberg EW, Borsboom GJ, van Houwelingen HC, Eijkemans MJ, Habbema JD. Validation and updating of predictive logistic regression models: a study on sample size and shrinkage. Stat Med. 2004 Aug 30;23(16):2567–86.

50. Riley RD, Lambert PC, Abo-Zaid G. Meta-analysis of individual participant data: rationale, conduct, and reporting. BMJ. 2010 Feb 5;340:c221.

